# Exposome Epidemiology for Suspect Environmental Chemical Exposures during Pregnancy Linked to Subsequent Breast Cancer Diagnosis

**DOI:** 10.1101/2023.06.20.23291648

**Authors:** Young-Mi Go, Jaclyn Weinberg, Sami Teeny, Piera Cirillo, Nickilou Krigbaum, Grant Singer, ViLinh Ly, Barbara Cohn, Dean P. Jones

## Abstract

Breast cancer is now the most common cancer globally, accounting for 12% of all new annual cancer cases worldwide. Despite epidemiologic studies having established a number of risk factors, knowledge of chemical exposure risks is limited to a relatively small number of chemicals. In this exposome research study, we used non-targeted, high-resolution mass spectrometry (HRMS) of pregnancy cohort biospecimens in the Child Health and Development Studies (CHDS) to test for associations with breast cancer identified via the California Cancer Registry. Second (T2) and third (T3) trimester archival samples were analyzed from 182 women who subsequently developed breast cancer and 384 randomly selected women who did not develop breast cancer. Environmental chemicals were annotated with the Toxin and Toxin-Target Database (T3DB) for chemical signals that were higher in breast cancer cases and used with an exposome epidemiology analytic framework to identify suspect chemicals and associated metabolic networks. Network and pathway enrichment analyses showed consistent linkage in both T2 and T3 to inflammation pathways, including linoleate, arachidonic acid and prostaglandins, and identified new suspect environmental chemicals associated with breast cancer, i.e., an N-substituted piperidine insecticide and a common commercial product, 2,4-dinitrophenol (DNP), linked to variations in amino acid and nucleotide pathways in T2 and benzo[a]carbazole and a benzoate derivative linked to glycan and amino sugar metabolism in T3. The results identify new suspect environmental chemical risk factors for breast cancer and provide an exposome epidemiology framework for discovery of suspect environmental chemicals and potential mechanistic associations with breast cancer.

## 1. Introduction

High-resolution metabolomics (HRM) uses advanced mass spectrometry and data science for non-targeted analysis of biological samples to obtain insight into an individual’s metabolism along with the individual’s exposome, including chemicals derived from diet, intestinal microbiome, dietary supplements, pharmaceuticals, personal use products and environmental exposures (Jones *et al*. 2012). The metabolome is a functional readout of the interactions of a person’s genes with exposures from diet and environment (Johnson *et al*. 2017; Jones *et al*. 2016; Niedzwiecki *et al*. 2019) and has been used extensively in combination with exposure measurements, such as air pollution (Liang *et al*. 2018; Ritz *et al*. 2022) and targeted environmental chemicals, such as DDT, PCBs and PBB (Hu *et al*. 2020; Walker *et al*. 2019), to gain understanding of functional associations with exposures. Use in a non-targeted manner for discovery of new chemical associations with disease, however, are limited. A vision for development of exposome epidemiology, i.e., using omics scale biomonitoring for disease epidemiology, is available (Jones and Cohn 2020). The current study was undertaken to apply exposome epidemiology concepts for discovery of potential new environmental factors contributing to breast cancer. The long-term goal for complex diseases such as breast cancer, which involves multiple genetic, behavioral and environmental factors, is to integrate omics scale biomonitoring with other factors to ultimately improve prediction and intervention strategies to decrease disease burden.

To perform a non-targeted exposome-wide association study of breast cancer, we used archival blood samples from the, Child Health and Development Studies (CHDS) cohort collected during pregnancy from 1959-1967. Second (T2) and third (T3) trimester archival samples, collected from 1959-1967, of 182 women who subsequently developed breast cancer, were compared to those from 384 women who did not develop breast cancer.

We developed an exposome epidemiology approach (**Fig 1**) in which we first performed non-targeted statistical analysis to select HRM features that were higher in cases than in non-cases. In this selection, we use raw *p*<0.05 as a cutoff for selection based upon the expectation that multiple environmental factors with small effect size may contribute to breast cancer, and this more liberal selection criteria is more appropriate for a small-population discovery study because it provides a better balance between Type 1 and Type 2 statistical error than more rigorous False Discovery Rate (FDR) criteria. We selected HRM features that were higher in cases than non-cases to search for potentially causative factors by matching to a xenobiotic database, Toxin and Toxin-Target Database (T3DB) (Lim *et al*. 2010; Wishart *et al*. 2015), recognizing that protective factors that are decreased in association with breast cancer would prioritize use of HRM features that were lower in cases and an alternative database, such as Food Database (FoodDB) (Scalbert *et al*. 2011). HRM features that were increased with breast cancer and annotated as environmental chemicals were then used for network analysis with the data-dependent community detection tool, xMWAS (Uppal *et al*. 2018). Pathway enrichment analysis of the most central communities was then performed with *mummichog* (Li *et al*. 2013), and targeted mass spectrometry was used to improve understanding of the annotated chemicals associated with breast cancer outcome. The results provide evidence for new candidate environmental breast carcinogens and information concerning possible mechanisms by which these agents contribute to breast cancer.

**Figure 1.**
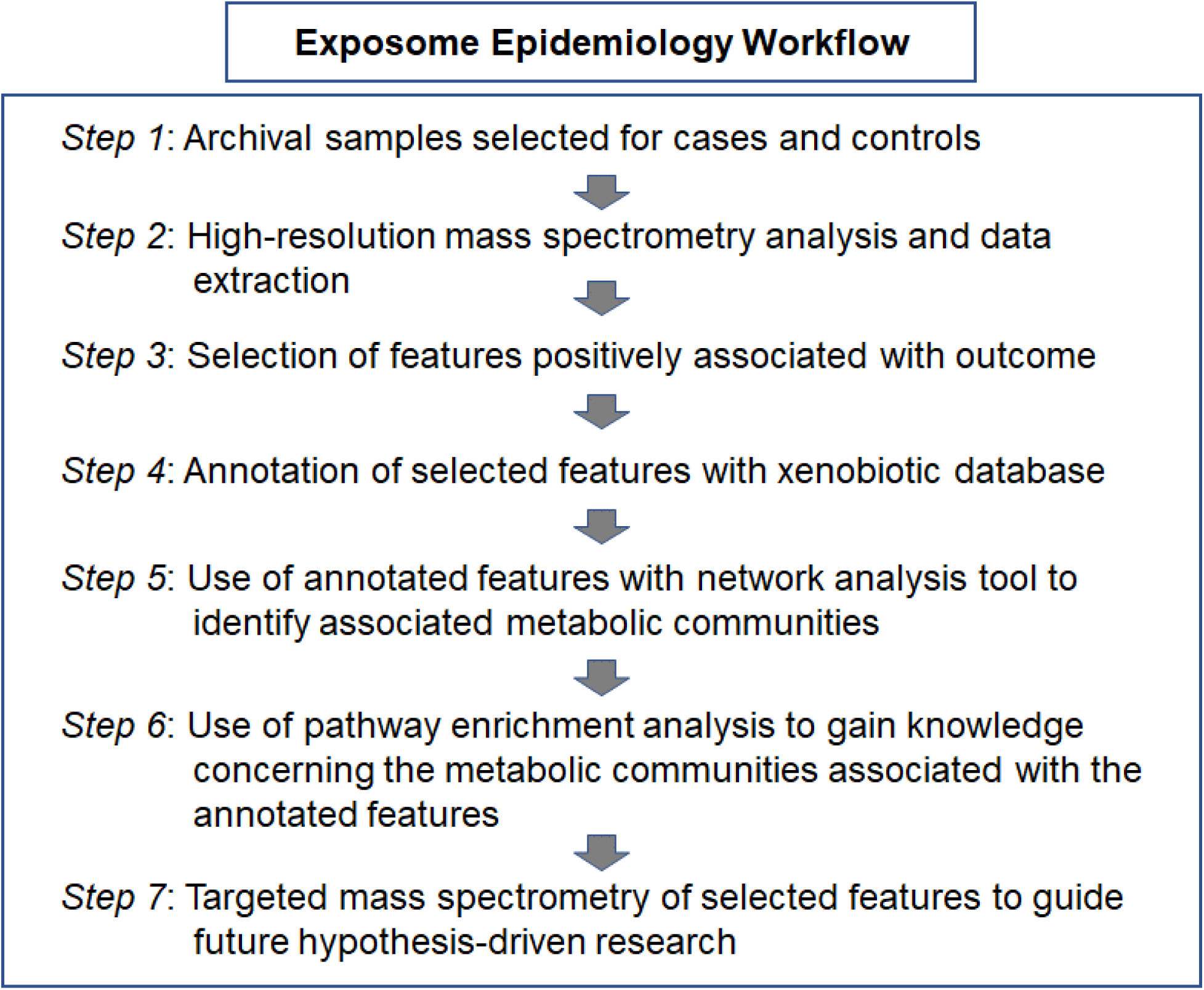
Exposome Epidemiology Workflow. The steps used in the current analysis are summarized in general form. In the first step, the study population and sample types can be selected for diverse purposes. In the second step, liquid chromatography, gas chromatography or other separation methods can be used with high-resolution mass spectrometry to generate tens of thousands of mass spectral features. For this purpose, multiple data extraction tools are available, and extraction parameters and filtration criteria can be selected to increase stringency or leniency in feature retention. For the third step, directionality of change is selected for features expected to have a causal or protective activity. For the fourth step, selected features are annotated using a relevant database. In the fifth step, selected and annotated features are used with network analysis to identify associated communities of mass spectral features. The sixth step applies pathway enrichment analysis to the associated communities of mass spectral features to characterize the associated biologic response. Finally, targeted mass spectral analysis of relevant mass spectral features provides prioritization for subsequent hypothesis-driven research to verify environmental chemical associations and underlying causal mechanisms.

## 2. Materials and methods

### 2.1 Samples and Assays

The CHDS recruited women residing in the Oakland and East Bay, California area who were members of the Kaiser Foundation Health Plan and received obstetric care for pregnancies between 1959 and 1966 with deliveries extending into 1967 (van den Berg *et al*. 1988). More than 98% of all eligible women enrolled.

Blood samples were collected from these mothers during pregnancy in each trimester and in the early post-partum period, processed to isolate serum, and stored since then at −20° Celsius. Second and third trimester archival samples available were analyzed for HRM and environmental chemicals. The samples used for the present study include second (T2, n=182)) and third (T3, n=172)) trimester archival samples of 201 women who subsequently developed breast cancer, compared to second (T2, n=384) and third (T3, n=351) trimester archival samples from 413 women who did not develop breast cancer. Breast cancer cases were identified by linkage to the California Cancer Registry for cases diagnosed through 1997. Record abstraction for cancer diagnoses to the California Cancer Registry is based primarily on pathology reports, and case identification is considered to be >99% complete after a 2-year lag (Perkins *et al*.). Cases were defined as mothers with incident invasive or noninvasive breast cancer diagnosed at a median age at diagnosis of 54 years (interquartile range, 13 years) with available prenatal serum and a standardized gross placental exam. Non-cases were an 8% sample of women not known to have breast cancer randomly selected among mothers with available prenatal serum and a standardized gross placental exam (Cohn *et al*. 2017). Breast cancer rates for included and excluded subsets in the CHDS cohort were highly comparable suggesting selection did not impose significant bias: 1.89 per 1,000 person-years (95% Confidence Interval ([95% CI] = 1.60, 2.23) for included vs. 1.88 per 1,000 (95% CI=1.59, 2.20) for excluded. It is possible, however, that we missed some cases of breast cancer, including among women who are identified in this study as “non-cases”. In this case, failure to identify cases among the non-case group would result in underestimating differences between cases and non-cases in these analyses and would not be expected to impact findings. The CHDS founding mothers voluntarily participated in an in-person interview and gave permission to access their own medical records and those of their children to researchers. The institutional review board of the Public Health Institute approved the present study (IRB#120-002), and we complied with all federal guidelines governing the use of human participants. Forty-six percent of cases and forty-five percent of the non-cases had available data and serum.

### 2.2 Chemicals

HPLC grade acetonitrile and methanol, LC-MS water and 98% formic acid were obtained from Sigma-Aldrich (St. Louis, MO). A mixture of 14 stable isotopic chemicals were used as an internal standard (Go *et al*. 2015) included [^13^C_6_]-D-glucose, [^15^N]-indole, [2-^15^N]-L-lysine dihydrochloride, [^13^C_5_]-L-glutamic acid, [^13^C_7_]-benzoic acid, [3,4-^13^C_2_]-cholesterol, [^15^N]-L-tyrosine, [trimethyl-^13^C_3_]-caffeine, [^15^N_2_]-uracil, [3,3-^13^C_2_]-cystine, [1,2-^13^C_2_]-palmitic acid, [^15^N,^13^C_5_]-L-methionine, [^15^N]-choline chloride and 2’-deoxyguanosine-^15^N ^13^C −5’-monophosphate from Cambridge Isotope Laboratories, Inc (Andover, PA).

### 2.3 High-resolution mass spectrometry

Serum samples were analyzed with liquid chromatography-high resolution mass spectrometry (LC-HRMS) as described previously (Jarrell *et al*. 2021; Jarrell *et al*. 2020). Briefly, 50 μL of serum was treated 2:1 (v/v) with acetonitrile, and 2.5 µL of the stable isotope standard mixture was added. Proteins were precipitated by incubation at 4°C for 30 min and removed by centrifugation for 10 min at 21000 x *g* at 4°C. Supernatants were placed in autosampler vials and maintained at 4°C in an autosampler until analysis. Two pooled human reference samples including NIST SRM1950 and Qstd [pooled plasma from 2 separate lots from Equitech-Bio, Inc (Kerrville, Texas)] were included. NIST SRM1950 was run at the beginning and end of the full sample set, and Qstd was included at the beginning and end of each batch of 20 samples.

Samples and reference materials were analyzed with three technical replicates using a High-Field Q-Exactive mass spectrometer (Thermo Fisher) with C18 chromatography and electrospray ionization (ESI) in negative mode. Data collection occurred continuously throughout 5 min of chromatographic separation from 85 to 1,275 mass-to-charge ratio (*m/z*). Data extraction was performed using *apLCMS* and *xMSanalyzer*, generating mass spectral features consisting of *m/z*, retention time (RT) and peak intensity. Feature and sample filtering retained features with a median CV of 50% or less, a minimum mean Pearson correlation coefficient of 0.7 between technical replicates of each sample, and presence in at least 30% of samples.

### 2.4 Selection of features associated with subsequent breast cancer

A Metabolome-Wide Association Study (MWAS) was used to identify *m/z* features associated with breast cancer outcome. Intensity values were log2 transformed, and LIMMA was performed using the R package, *xmsPANDA* (https://github.com/kuppal2/xmsPANDA), with retention of features at raw *p*<0.05.

### 2.5 Annotation of Mass Spectral Features and Metabolite Identification

To select mass spectral features of environmental interest, features positively associated with subsequent breast cancer diagnosis were subjected to a multistage clustering algorithm and annotated with xMSannotator (Uppal *et al*. 2017) using T3DB (Lim *et al*. 2010; Wishart 2015) at 5 ppm tolerance. For selected chemicals, identities were confirmed when possible by accurate *m/z* match, co-elution with authentic standards and ion dissociation mass spectrometry (MS/MS); Level 1 identification by criteria of Schymanski et al (Schymanski *et al*. 2014)].

### 2.6 Targeted mass spectrometry analysis in identification of features

Annotated chemicals at the center of the *xMWAS* network structures underwent additional investigation by ion fragmentation, structure analysis, and spectral matching to publicly available spectral libraries. Plasma samples with the highest intensities of the chemicals of interest were selected and analyzed with C18 liquid chromatography (Dionex Ultimate 3000) and MS^2^ ion fragmentation (Thermo Scientific Fusion) with negative ESI. The mass spectrometer was set to scan a minimal range of *m/z*’s at 60k resolution for MS^1^. Then, an inclusion list isolated the *m/z* of interest. MS^2^ fragmentation was acquired in HCD mode with a normalized collision energy of 30% and analyzed at 30,000 resolution using the dual-pressure linear ion trap. Raw MS^1^ and MS^2^ spectra were analyzed in xCalibur QualBrowser (Thermo). Fragmentation patterns were analyzed with ChemDraw (PerkinElmer Informatics) and compared against experimental spectra in the MassBank of North America (MoNA) library. MoNA MS^2^ spectra were selected for negative ESI and similar collision energy. When MoNA spectra were not available, experimental spectra were searched against the NIST Tandem Mass Spectral Library, which provides a spectral match percentage but no information on collision energy or ionization polarity. To align with the goal of high-throughput analyses, chromatographic gradients and fragmentation settings were maintained to be consistent with the original analysis and not extensively optimized for each chemical. Under these conditions, the complexity of the plasma sample matrix and the low abundance of environmental chemicals does not allow for the isolation of one precursor and its fragments. Therefore, in some spectra, the precursor mass is easily visible within 10 ppm error, but the fragments are not clearly visible due to interference from other ions. Spectral peaks corresponding to fragments were identified by their theoretical structures in “ChemDraw” with unit resolution in the dual-pressure linear ion trap detector.

### 2.7 Network and Pathway Analyses

Annotated environmental chemicals were tested for associations with *m/z* features using *xMWAS* based on partial least-squares regression (Uppal *et al*. 2018). Thresholds for inclusion in the networks were |r| > 0.30 and *p* < 0.05. Pairwise results from MWAS of the metabolites used for generation of network structures were used for pathway enrichment analysis using *mummichog (v3)* (Li *et al*. 2013). Enriched pathways were filtered for those that included at least 3 significantly associated metabolites at *p* < 0.05.

## 3. RESULTS

### 3.1 Study population demographics

T2 and T3 samples were available from 201 individuals who subsequently developed breast cancer. These individuals had a median age of 31 y at time of blood collection, and the individuals without breast cancer had a median age of 36 (p<0.0001). Serum collection occurred between 1960 and 1964, with T2 blood collection occurring at a median gestational age of 161 and 160 days in cases and non-cases, respectively. For T3, blood collections were at a median gestational age of 249 and 250 days, respectively. Characteristics of cases and non-cases are provided in **Table 1**.

**Table 1.** Characteristics of Study Population

| **Characteristic** | **Non-Cases**<br>**(N=413)** |  | **Cases (N=201)** |  |
| --- | --- | --- | --- | --- |
|  | **Median** | **(IQR)** | **Median** | **(IQR)** |
| Year of mother's birth | 1936 | (9) | 1931 | (11) |
| Year of blood draw | 1962 | (2) | 1962 | (2) |
| Gestational day of T2 blood draw | 160 | (33) | 161 | (29) |
| Gestational day of T3 blood draw | 250 | (17) | 249 | (16) |
| BMI at first prenatal visit (kg/m<sup>2</sup>)<sup>a</sup> | 22.1 | (4.3) | 22.6 | (4.2) |
| **Characteristic** | **Percent** |  | **Percent** |  |
| Race |  |  |  |  |
| non-Hispanic Caucasian | 70 |  | 69 |  |
| African American | 20 |  | 19 |  |
| Hispanic | 2 |  | 1 |  |
| Asian | 6 |  | 7 |  |
| Mixed | 2 |  | 3 |  |

### 3.2 Metabolic feature profiling on T2 and T3 serum

To select *m/z* features which were positively associated with subsequent breast cancer diagnosis, we retained 9042 *m/z* features present in at least 80% of samples for statistical analyses and selection of features for annotation. For T2 and T3 measurements, no features had FDR<0.05 and therefore none are likely to be useful biomarkers for breast cancer risk. Any of the *m/z* features selected with raw p<0.05 could contribute to breast cancer risk, and 521 features had p<0.05 for T2, and 557 features had p<0.05 for T3. Of these *m/z* features, 188 were higher in breast cancer than non-cases in T2 samples while 151 were higher in breast cancer than non-cases in T3 samples (**Table 2**). These positively associated *m/z* features were selected to search for possible matches to environmental chemicals.

**Table 2.** HRM profiling of T2 and T3 serum samples comparing breast case and non-case groups (*P*≤0.05). BC, breast cancer.

|  | T2 (182 BC, 384 non-cases) | T3 (172 BC, 351 non-cases) |
| --- | --- | --- |
| Total number of metabolic features | 9042 | 9042 |
| Features differing at. $p < 0.05$ | 521 | 557 |
| Differing features with BC/non-cases ratio $> 1.0$ | 188 | 151 |

### 3.3 Environmental chemical annotation of suspect chemicals in breast cancer group

Most of the selected *m/z* features are likely to represent endogenous metabolites, but environmental chemicals contributing to subsequent breast cancer could also be present. To search for suspect environmental chemicals associated with subsequent breast cancer diagnosis, we used xMSannotator, a network-based computational tool, with the Toxin and Toxin-Target Database, T3DB (Wishart 2015; Wishart *et al*. 2007), to further characterize the mass spectral features which were higher in their intensities in women who went on to develop breast cancer. xMSannotator uses a multistage clustering algorithm in which intensity profiles, retention time characteristics, mass defect, and isotope/adduct patterns are used to assign confidence levels to annotation results relative to publicly available databases, such as T3DB. The annotated environmental chemicals for T2 and T3 are shown in **Table 3** and **Table 4**, respectively. More environmental chemicals were annotated in T2 than T3 serum, yielding 17 and 7, respectively, and these included matches to chemicals widely used for anti-inflammatory, anticonvulsant and antipsychotic drugs, quaternary ammonium salt, pesticides, herbicides, fungicides, plasticizers, preservatives, cleaning materials, and flavorings. Importantly, at this level of investigation, these annotations are only suspect chemicals and require further investigation and verification (see below). The magnitude of difference was between 10% and 30% higher in women who went on to develop breast cancer compared to those who did not (p ≤ 0.05, **Tables 3 and 4**).

**Table 3.**
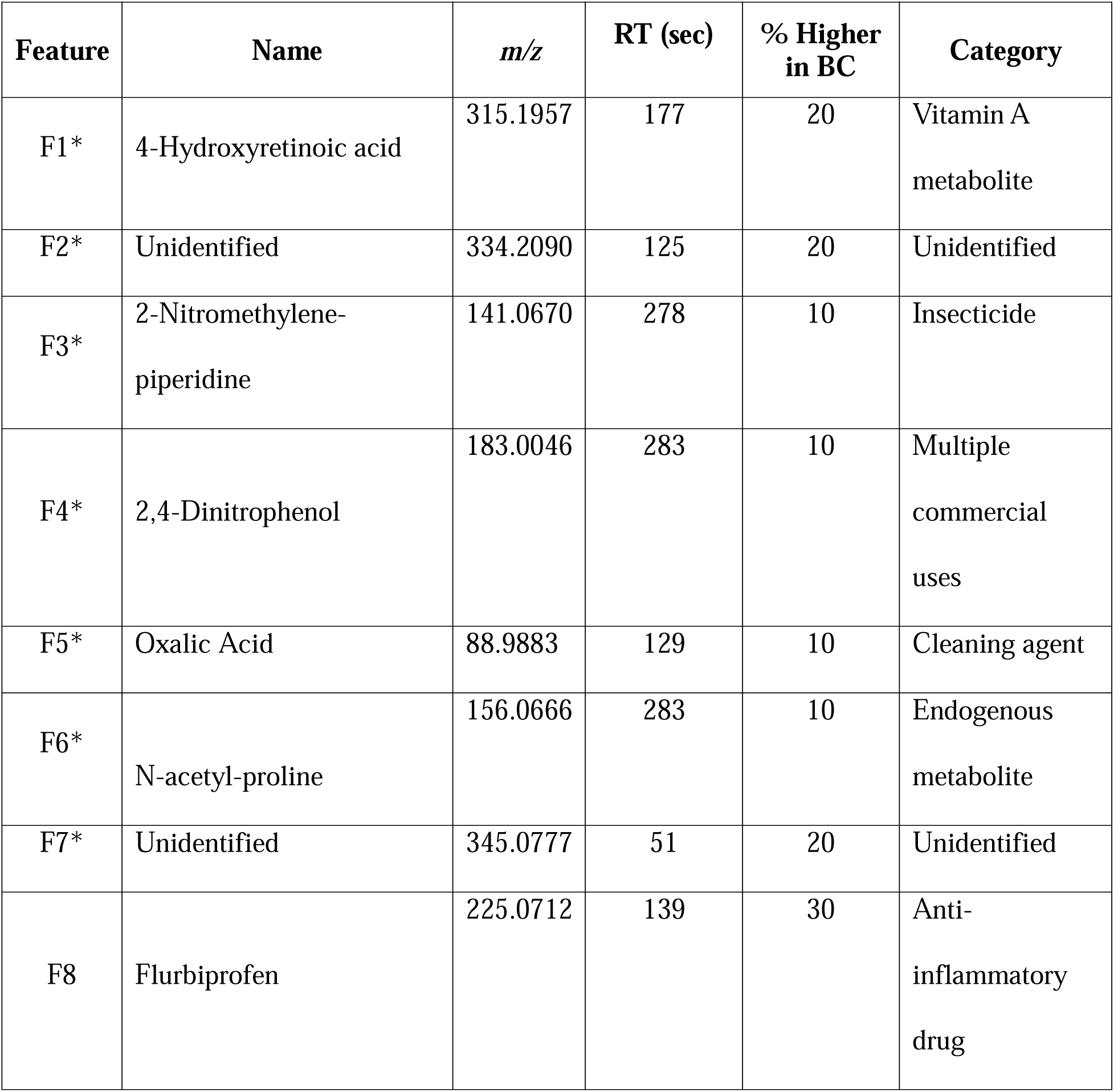
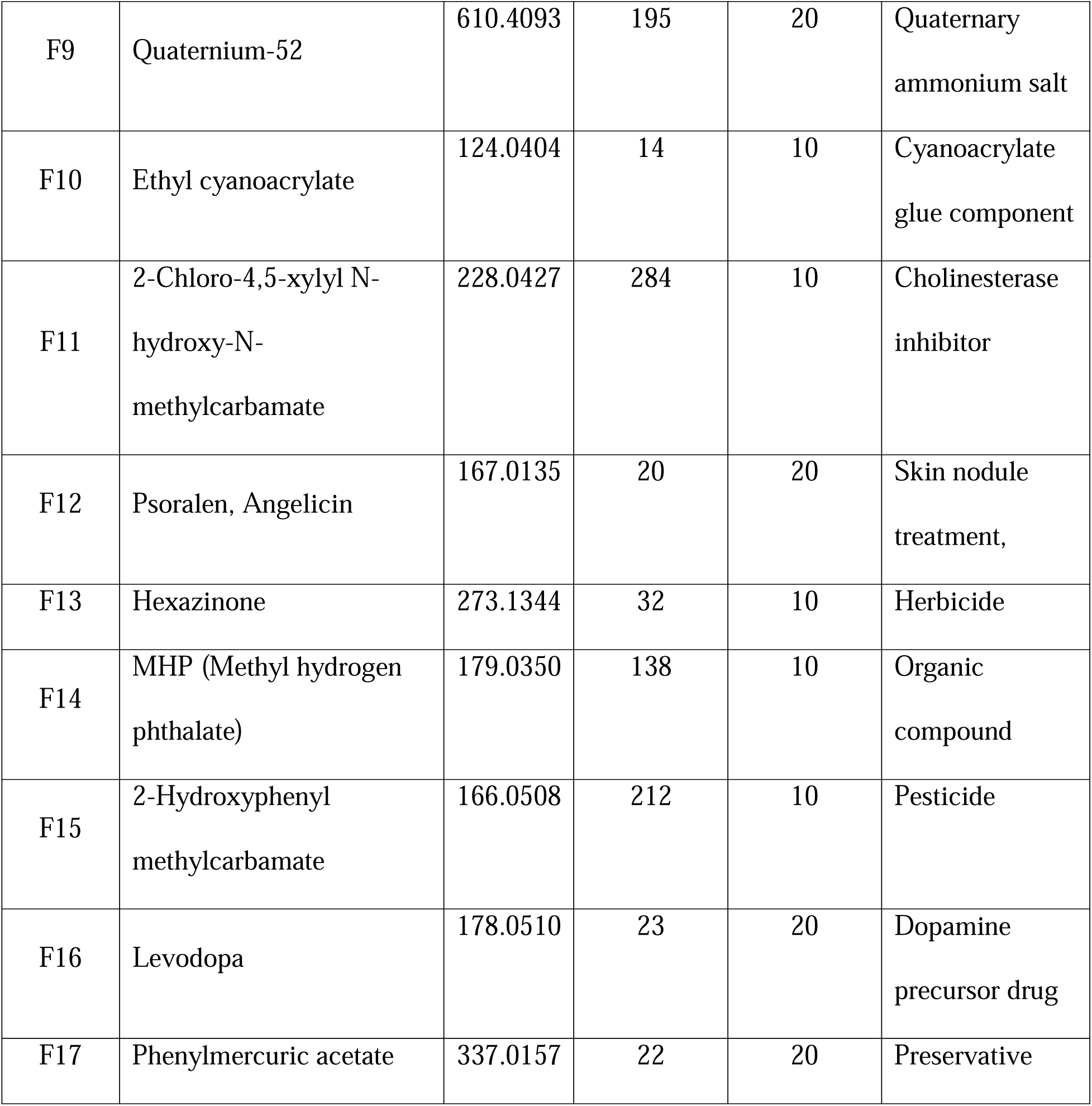
T3DB-annotated *m/z* features higher in T2 serum of women who subsequently developed breast cancer (BC) compared with non-cases (*P* ≤ 0.05). Targeted mass spectrometry analysis was performed to aid in identification of features F1 to F7 which were found to occur in central communities of Fig 2, indicated by asterisk (*). T3DB annotations are provided for others (F8-F17).

**Table 4.**
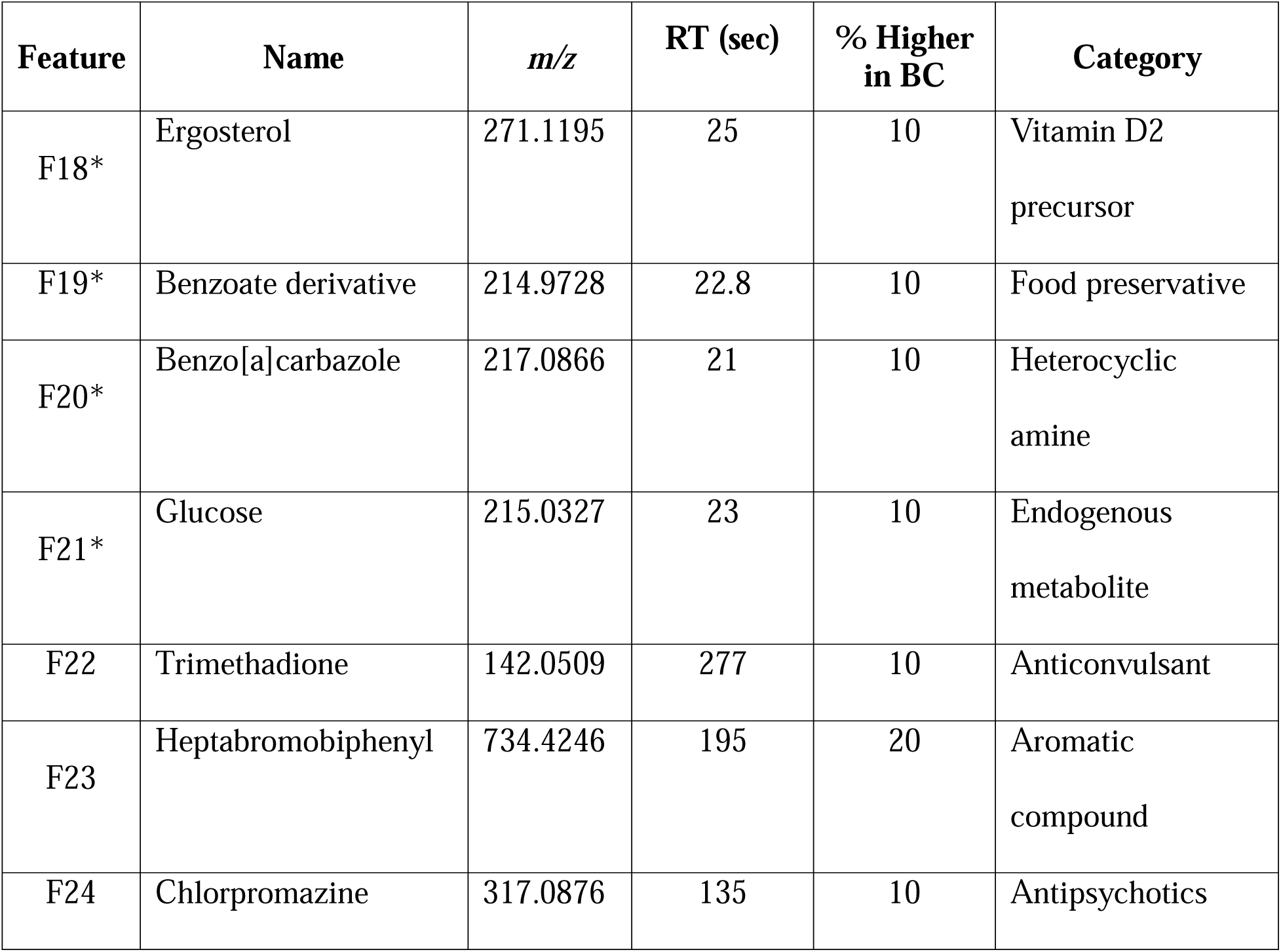
T3DB-annotated *m/z* features higher in T3 serum of women who subsequently developed breast cancer (BC) compared with those in the non-case women (*P* ≤ 0.05). Targeted mass spectrometry analysis was performed to aid in identification of features F18-F21 which were found to occur in central communities of Fig 4, indicated by asterisk (*). T3DB annotation is provided for others (F22-F24).

| Feature | Name | $m/z$ | RT (sec) | % Higher in BC | Category |
| --- | --- | --- | --- | --- | --- |
| F18* | Ergosterol | 271.1195 | 25 | 10 | Vitamin D2 precursor |
| F19* | Benzoate derivative | 214.9728 | 22.8 | 10 | Food preservative |
| F20* | Benzo[a]carbazole | 217.0866 | 21 | 10 | Heterocyclic amine |
| F21* | Glucose | 215.0327 | 23 | 10 | Endogenous metabolite |
| F22 | Trimethadione | 142.0509 | 277 | 10 | Anticonvulsant |
| F23 | Heptabromobiphenyl | 734.4246 | 195 | 20 | Aromatic compound |
| F24 | Chlorpromazine | 317.0876 | 135 | 10 | Antipsychotics |

### 3.4 Metabolome-Wide Association Study (MWAS) of T2 Environmental Chemicals

Prior studies have identified inflammatory lipid pathways (linoleate, arachidonate, prostaglandin), lipid and energy metabolism pathways (fatty acids, TCA cycle), oxidative stress pathways (methionine and cysteine), and nitrogen metabolism pathways (urea cycle, pyrimidine, purine) as top metabolic pathway associations with environmental chemicals and breast cancer (Hu *et al*. 2019; Li *et al*. 2020; Walker *et al*. 2019). To determine whether the features annotated as environmental chemicals (**Table 3**) are associated with changes in metabolites and metabolic pathways, we used data-driven integration and network analysis of the T3DB annotated features and all 9042 extracted metabolic signals, with xMWAS (Uppal *et al*. 2018). As shown in **Fig 2**, network analysis showed three metabolic communities, Community 1 (C1, orange), Community 2 (C2, blue) and Community 3 (C3, green). C1 was positively associated with *m/z* 315.1957 (F1, **Fig 2**, **Table 3**). We performed metabolic pathway enrichment analysis (**Fig 3**) on the 716 metabolic features associated with C1 using the *mummichog 3* software (Li et al. 2013). Associated pathways included linoleate, arachidonate and prostaglandins (**Fig 3A**), which are closely related to inflammation and oxidative stress. Database searches showed that *m/z* 315.1957 matched multiple lipids (e.g., prostaglandins, resolvins), consistent with the strong associations with related lipid species having activities in inflammation. The vitamin A pathway was also associated with C1 (**Fig 3A**), and targeted mass spectrometry with MS/MS analysis showed a likelihood that *m/z* 315.1957 was 4-hydroxyretinoic acid (**F1, Supplemental Fig. S1**).

**Figure 2.**
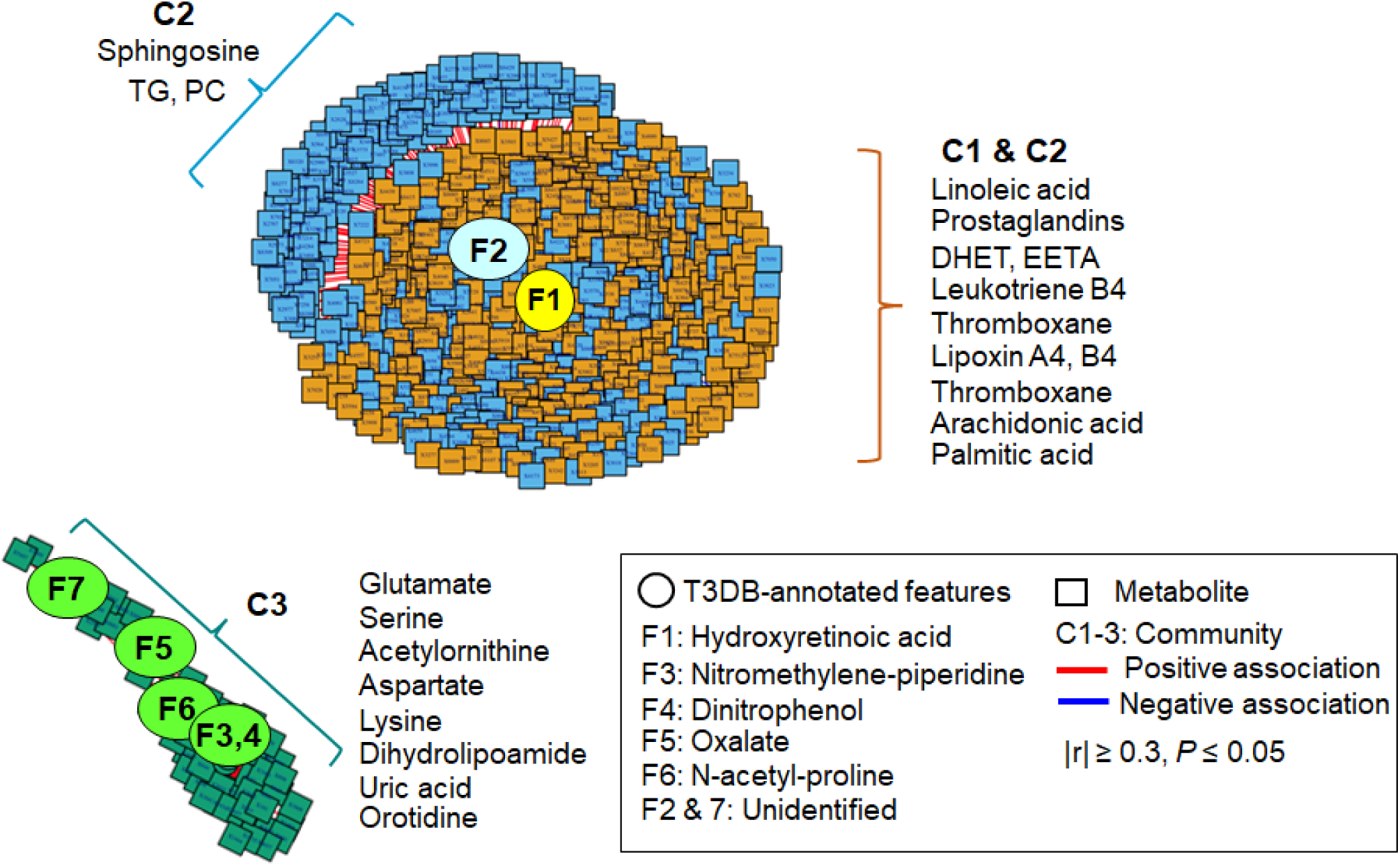
Metabolome-Wide Association Study (MWAS) of T3DB-annotated chemicals higher in second trimester (T2) serum. Association of seventeen T3DB-annotated features (Table 3) that are higher in breast cancer than non-cases with the metabolome (9,042 metabolic features) from T2 serum (n=384 for non-cases, n=182 for breast cancer) are examined using xMWAS. The outcome of xMWAS analysis is visualized with two separate networks at |r| ≥ 0.3. The networks include three communities (C1, C2, C3) and show central seven chemicals (F1-F7) with tight connections [C1 (orange). C2 (blue), C3 (green)]. Representative metabolites of pathways associated with community are shown next to each community. DHET: dihydroxyeicosatrienoic acid, EETA: epoxyeicosatrienoic acid, TG: triglyceride, PC: phosphatidylcholine.

**Figure 3.**
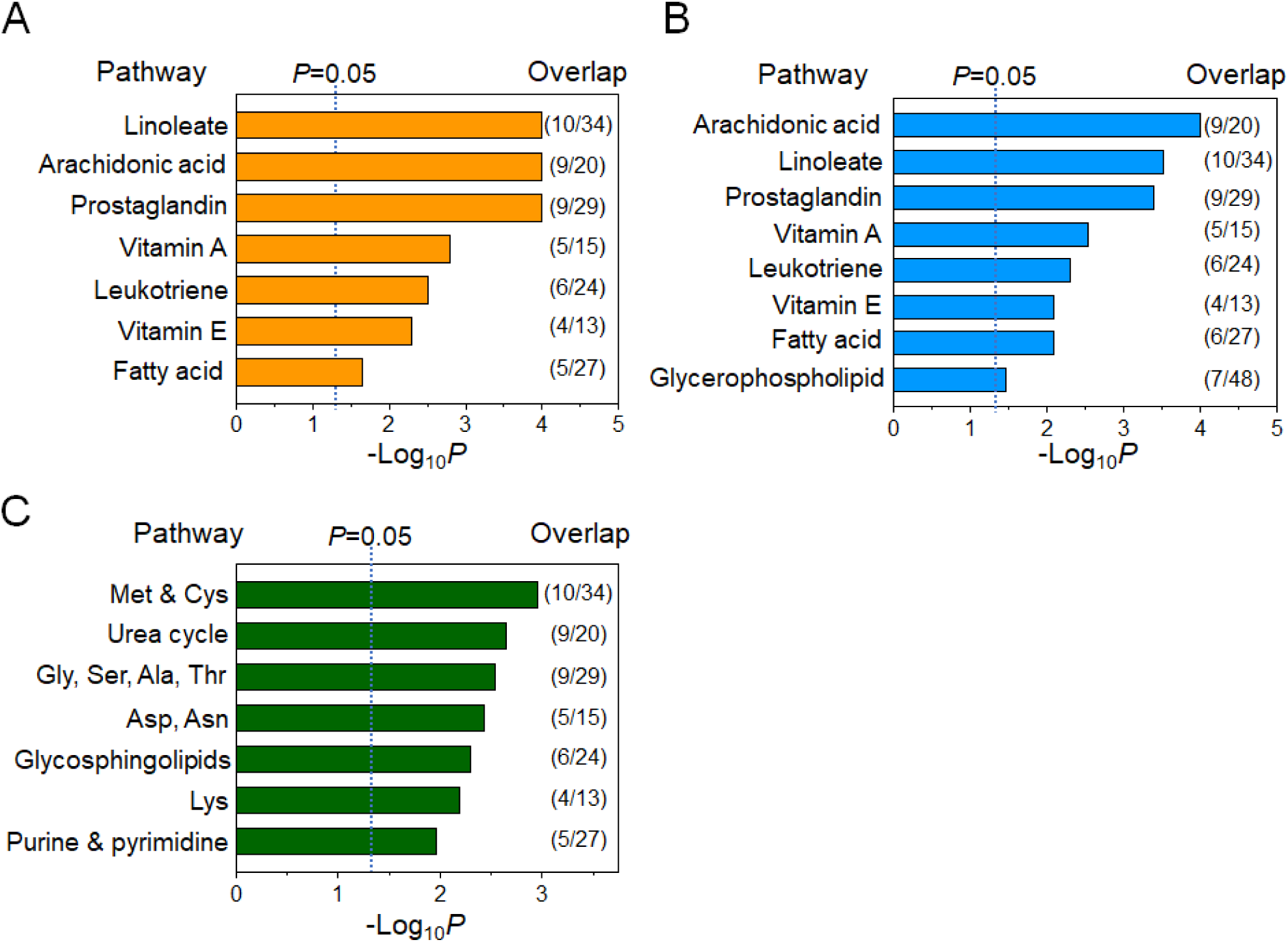
Metabolic pathway associated with second trimester (T2) chemicals. Pathway enrichment analysis with metabolites of three network communities in Fig. 2 was conducted using mummichog. **A)** A total of seven pathways were found altered with *m/z* 315.1957 (F1) in C1, **B**) Eight pathways were altered with *m/z* 334.2090 (F2) in C2, and **C)** Seven pathways were altered with environmental chemicals and drug metabolites (F3-F7) (p < 0.05). The ratio of selected metabolites mapped to the listed pathway over the number of total pathway metabolites detected is provided to the right of each bar.

C1 extensively overlapped with Community 2 (C2, blue), which was positively associated with *m/z* 334.2090 (F2, **Fig 2**, **Table 3**). In addition to pathway associations with prostaglandins, linoleic acid and arachidonic acid, a subset of C2 included phospholipids, e.g., sphingosine, phosphatidyl choline (PC), and triglyceride (blue, **Fig 2**), associated with glycerophospholipid metabolism. Thus, this chemical defines a separate cluster of metabolites associated with breast cancer than those in C1, but this feature remains unidentified because subsequent mass spectrometry analysis failed to provide support for identification. Together, results from C1 and C2 support prior findings that inflammatory lipid and other lipid pathways associated with DDT, PCB and PBB are associated with breast cancer, but do not reveal any new suspect environmental chemicals.

A third community, C3, had metabolomic associations (green) which were separated from C1 and C2 and positively associated with *m/z* features annotated as environmental chemicals, including an insecticide (F3, nitromethylene-piperidine), a wood preservation and dye production chemical (F4, 2,4-dinitrophenol), oxalate (F5), and F6, *m/z* 156.0666 and F7, *m/z* 345.0777 (**Fig 2**, **Table 3**). Of these, oxalate was previously confirmed, and supportive targeted mass spectrometry data was obtained for 2-nitromethylene-piperidine and 2,4-dinitrophenol (**F3 and F5, Supplemental Fig. S1**). The *m/z* feature 156.0666 (F6) was present at high concentrations in most samples and appeared likely to be an endogenous metabolite, N-acetylproline (**F6, Supplemental Fig. S1**). No supportive mass spectrometry data could be obtained for the *m/z* feature 345.0777 (F7). The 56 metabolic features in C3 that were positively associated with these annotated environmental chemicals were related to pathways for antioxidant methionine (Met) and cysteine (Cys) regulation, urea cycle, amino acids (glycine, serine, alanine, threonine, aspartate, asparagine, lysine), glycerophospholipids, and purine and pyrimidine nucleotide metabolism (**Fig 3C**). Representative metabolites associated with these pathways of each community are indicated next to each community (**Fig 2**) and included amino acids (glutamate, serine, lysine, aspartate), a pyrimidine (orotidine), a purine (uric acid), and a coenzyme for mitochondrial electron transfer and the TCA cycle (dihydrolipoamide). Collectively, the results for T2 serum of women who subsequently developed breast cancer show that two suspect environmental chemicals, nitromethylene-piperidine and 2,4-dinitrophenol, along with oxalate, are closely associated with metabolic perturbations related to amino acid and nucleotide metabolism. Other communities included lipids functioning in inflammation, sphingolipids and lipids, but no suspect environmental chemicals were identified for these communities.

### 3.5 MWAS of T3 Environmental Chemicals and alterations in metabolic pathway

Following the same experimental approach shown above, we examined the relationship of annotated matches of T3 suspect environmental chemicals (**Table 4**) and changes in metabolites using xMWAS (Uppal *et al*. 2018). Two major metabolic communities were identified (**Fig 4**), with the first community (labeled C4, orange) including metabolites associated exclusively with *m/z* 271.1195 (F18) and second community (C5, blue) including metabolites associated with *m/z* 214.9728 (F19), *m/z* 217.0866 (F20) and *m/z* 215.0327 (F21) (**Fig 4**). Targeted mass spectrometry analysis of *m/z* 271.1195 (F18) showed that this feature was likely ergosterol or related derivative (**F18, Supplemental Fig. S2**). Pathway enrichment analysis of the 650 metabolic features of C4 showed associated pathways for arachidonate, leukotriene, linoleate, vitamin A, prostaglandins, vitamin E and steroid hormones (**Fig 5A**). Most of these pathways are closely related to inflammation and oxidative stress, as also found in T2 associations in C1 and C2 (**Fig 3A, 3B**). Targeted mass spectrometry analyses showed that *m/z* 214.9713 (F19) was likely a bromine adduct of a benzoic acid metabolite (**F19, Supplemental Fig. S2**); *m/z* 217.0866 (F20) had an MS^2^ spectrum with 83% match to benzo[a]carbazole (**F20, Supplemental Fig. S2**); *m/z* 215.0327 (F21) was a high abundance signal from glucose (**F21, Supplemental Fig. S2**). Pathway enrichment analysis of the 155 metabolic features in C5 showed associations for sialic acid, pentose phosphate, N-glycan degradation, galactose, phosphatidylinositol, glycolysis and gluconeogenesis, butanoate, and glycosphingolipid pathways (**Fig 5B**). Representative metabolites associated with these pathways are indicated next to each community (**Fig 4**). Collectively, suspect environmental chemicals in Communities 4 and 5 include a benzoate derivative and benzo[a]carbazole, and metabolic pathway associations suggesting that breast cancer effects could be mediated through inflammatory lipids, glucose-related metabolism and estrogenic signaling.

**Figure 4.**
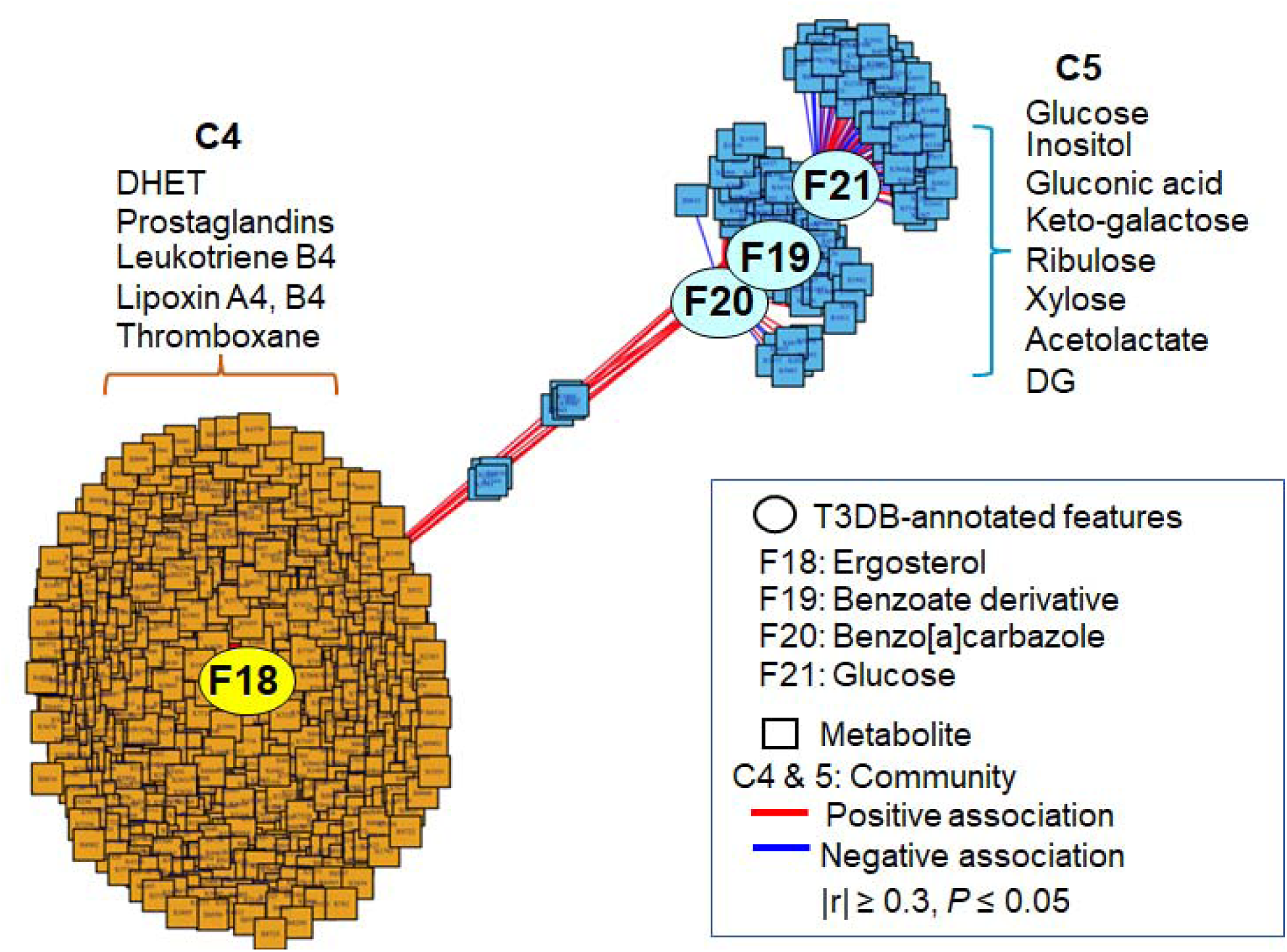
Metabolome-Wide Association Study (MWAS) of T3DB-annotated chemicals higher in third trimester (T3) serum. Association of seven T3DB-annotated chemicals (Table 4) that are higher in breast cancer than non-cases with the metabolome (9,042 metabolic features) from T3 serum (n=351 for non-cases, n=172 for breast cancer) are examined using xMWAS. The network is visualized at |r| ≥ 0.3 and includes two communities (C4 and C5) showing central four chemicals with tight connections [C4 (orange)] than C5 (blue)]. Representative metabolites of pathways associated with community are shown next to each community.

**Figure 5.**
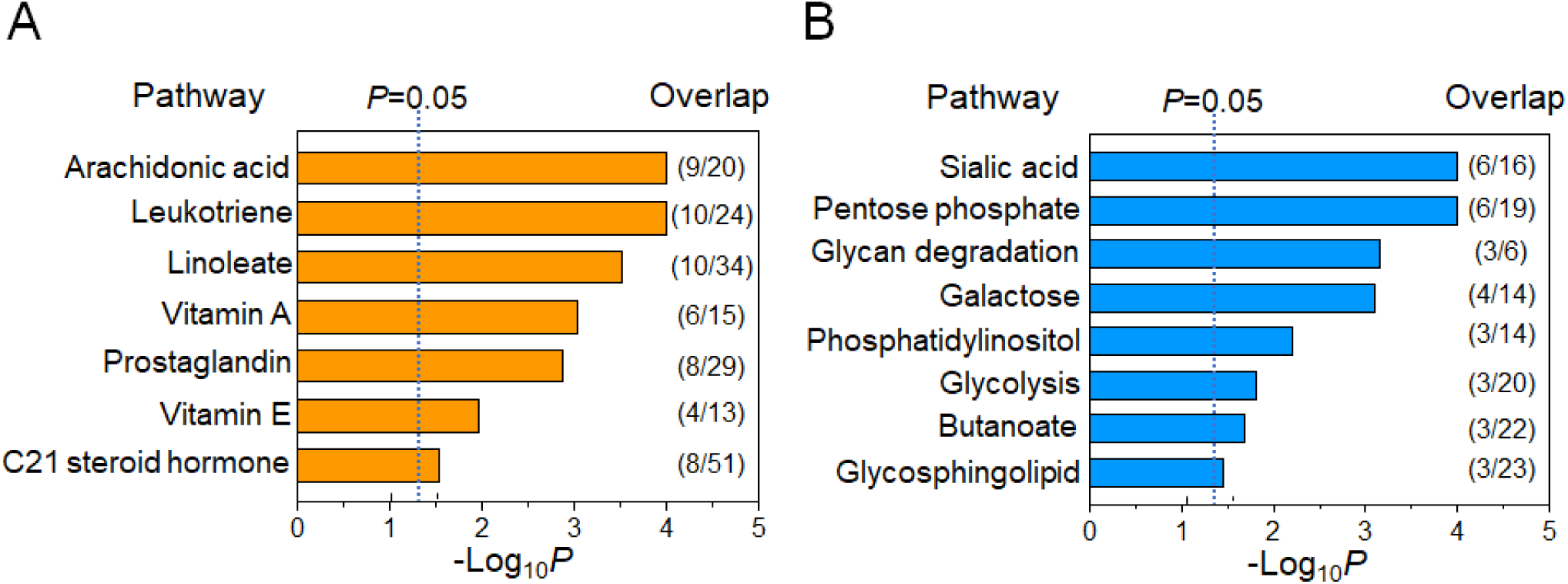
Metabolic pathway associated with T3 chemicals. Pathway enrichment analysis with metabolites of two network communities in Fig. 4 was conducted using mummichog. **A)** Seven pathways were found altered with F18 chemical in C1and **B**) eight pathways were altered with chemicals (F19-F21) in C2 (p < 0.05). The ratio of selected metabolites mapped to the listed pathway over the number of total pathway metabolites detected is provided to the right of each bar.

### 3.6 Comparisons of T2 and T3 suspect chemical-metabolic pathway associations

In comparison of findings for T3 and T2, metabolic pathways for C4 in T3 showed considerable overlap with the pathways associated with C1 and C2 in T2. In contrast, the remaining community, C5, in T3 differed substantially from the remaining community, C3, in T2. Specifically, in T3, we identified a suspect benzoate and benzo[a]carbazole in breast cancer cases that were associated with pathways of carbohydrate metabolism functioning in extracellular matrix turnover and complex carbohydrate metabolism. In T2, we identified a suspect insecticide, 2-nitromethylpiperidine, a widely used commercial chemical, 2,4-dinitrophenol, and oxalic acid, in breast cancer cases that were associated with central pathways functioning in amino acid homeostasis, urea cycle, pyrimidine and purine metabolism, and defense against oxidative stress.

## 4. Discussion

The present study applies a new exposome epidemiology approach to discover suspect environmental chemicals that may be implicated in breast cancer development. New suspect chemicals from this analysis include an insecticide, nitromethylene-piperidine; a common commercial chemical, 2,4-dinitrophenol; a heterocyclic amine with diverse commercial applications, benzo[a]carbazole; a benzoate derivative which could be derived from natural products or environmental chemicals; and oxalate, an endogenous metabolite that is also used as a cleaning agent. These chemicals were at higher abundance in serum of women decades before breast cancer diagnosis. Importantly, the network analysis is data-dependent, raising the possibility that individual chemicals could have a causal role in breast cancer or that breast cancer could occur through interaction of multiple chemicals in causal mechanisms. In the latter case, individual variations in exposure could have important impact on risk.

New approaches to identify suspect carcinogens are critical because research over the last 50 years has not translated to a strategy for individual breast cancer prevention. The suspect chemical signal F18 (*m/z* 271.1195) in Community 4, was consistent with identification as ergosterol, which is derived from yeast and precursor of vitamin D2. Pathway mapping of this community showed overlap with inflammatory lipid pathways found in Communities 1 and 2, and also included vitamin A metabolism, which is consistent with feature F1 in Community 1, 4-hydroxyretinoic acid, being a suspect chemical. Many diet-derived carotenoids can be converted to isobaric species to 4-hydroxyretinoic acid, and these are not distinguished by the HRM methods used. Thus, future research will be needed to address the potential for diet-environment interactions, such as implied by the overlap of estrogen-, vitamin D- and vitamin A-linked metabolites. Pregnancy impacts initiation, progression, and susceptibility to breast cancer (Troisi *et al*. 2018), and therefore pregnancy provides an appropriate time frame for study. Later age at pregnancy is a long-established breast cancer risk factor (Albrektsen *et al*. 2005; MacMahon *et al*. 1970) which has become increasingly common and is still unexplained. Because first birth rates have increased 6-fold for women ages 35-39 from 1973-2006 (Martin *et al*. 2015), more detailed understanding of the respective contributions and interactions must be a priority to learn how to mitigate risk for this population group of higher risk women.

In T2, an additional chemical cluster containing an insecticide (nitromethylene-piperidine), a cleaning agent (oxalate), a chemical used in wood preservation and dye production (dinitrophenol), and two unidentified chemical features (F2, F7), were associated with multiple amino acid pathways and nitrogen metabolism (urea cycle, pyrimidine, purine). The amino acid pathways overlap with pathways previously found to vary with persistent organic pollutants, *p,p’*-DDT (Hu *et al*. 2020) and PBBs (Walker *et al*. 2019). In a multigenerational study, *p,p’*-DDT exposure in women before puberty was found to be associated with breast cancer in mothers (Cohn *et al*. 2007) and in utero *o’,p’*-DDT exposure was associated with breast cancer in daughters (Cohn *et al*. 2015). Polybrominated biphenyls (PBB) are also persistent organic pollutants that cause breast cancer (IARC 2015). Potential mechanistic connections between dintrophenol, nitromethylene-piperidine, oxalate, and these POPs are not apparent, and mechanisms by which POPs disrupt central amino acid and nitrogen metabolism pathways are not known. In particular, amino acids are essential for growth and development, and disruption of pyrimidine and purine metabolism can be expected to have long-term consequences on metabolic programming which could contribute to cancer susceptibility. Thus, these findings warrant further investigation into underlying mechanisms related to breast cancer development.

In T3, the large metabolic community (C4, Fig 4) containing inflammatory lipids was linked to a sparse community (C5, Fig 4) containing glucose, benzo[a]carbazole and a suspect benzoate derivative. Of potential importance in this network, the glucose signal was negatively associated with metabolites connected to sialic acid, N-glycan and other pathways associated with extracellular matrix and turnover of connective tissue. Benzoate is conjugated with glycine for elimination, and the role of glycine in folate-dependent metabolism through the vitamin B_6_-dependent hydroxymethyltransferase raises the possibility that benzoate derivatives could have unrecognized pathogenic effects. Similarly, the heterocyclic amine, benzo[a]carbazole, could be bioactivated by Cyp enzymes to generate mutagenic species which have not been characterized. Future hypothesis-driven research will be needed to address these possibilities.

The exposome epidemiology approach used in the present study has important assumptions, i.e., that chemical exposures which increase cancer risk can occur decades before breast cancer occurrence, that these exposures are detectable and at higher abundance in serum decades prior to breast cancer detection, and that network analyses of HRM data are sufficient to detect these exposures and link them to biologic responses. With these assumptions, all HRM features from archival blood serum of women collected decades before breast cancer diagnosis can be used in a non-targeted manner to select ones that are increased (p<0.05) in association with breast cancer occurrence. This selection criterion is not suitable for biomarker development because of multiple testing; however, this cutoff is suitable for discovery of potential chemical risk factors because any of the features at this cutoff could be correct. The selected HRM features are then used with a toxic exposome database, T3DB, to obtain accurate mass matches to known toxic chemicals, and these are subjected to non-targeted network and pathway enrichment analyses to identify environmental chemical-metabolic network associations linked to breast cancer outcome.

Exposome epidemiology will benefit from populations of tens or hundreds of thousands of individuals by enabling detection of exposures impacting small numbers of women and/or having only small contributions individually to breast cancer risk. The present study with only hundreds rather than thousands of individuals has limited capability to detect carcinogens with small effect size. Additionally, selection of HRM features for statistical selection in the current study required that features be present in 80% of the samples. Environmental chemicals and personal use products such as hair dyes cannot be expected to be present in 80% of the women and unlikely to be detectable. Application of untargeted gas chromatography-high-resolution mass spectrometry, which provides capabilities to measure other hydrophobic and volatile environmental chemicals (Hu *et al*. 2021), can be used to enhance coverage of environmental chemicals. Thus, future studies will benefit from larger population sizes, inclusion of complementary chemical analyses, and use of statistical methods which can accommodate sparse environmental chemical detection.

## 5. Conclusion

Recent technological and statistical advances in high-resolution metabolomics (HRM) provide capabilities for omics scale biomonitoring of chemicals derived from the environment along with endogenous metabolism and chemicals from the diet, intestinal microbiome, dietary supplements, pharmaceuticals, and personal use products. Because the metabolome is a functional readout of the interactions of a person’s genes with exposures, HRM of biologic samples provides one of the most accessible ways to connect environmental exposures with biologic status to anticipate breast cancer. As shown in the present study, these can be used with network and pathway analysis to identify suspect environmental chemicals and functional communities linked to breast cancer outcome. Such network approaches can be broadly applied to discover how life-long exposures impact personal cancer risks. For breast cancer, we believe that this approach will yield critically needed protocols to enhance protection or mediate pregnancy-associated risk.

## Data Availability

All data produced in the present study are available upon reasonable request to the authors

## Abbreviations

BC: breast cancer
FDR: false discovery rate
HRM: high-resolution metabolomics
HRMS: high-resolution mass spectrometry
LC-HRMS: liquid chromatography-high resolution mass spectrometry
MWAS: metabolome-wide association study
*m/z*: mass to charge
RT: retention time
T2: second trimester
T3: third trimester
T3DB: the toxin and toxin-targeted database

## Funding

This study was supported by Department of Defense Grant W81XWH2010103, and National Institute of Environmental Health and Science grant R21 ES031824, R01 ES031980 and P30 ES019776.

## Notes

### Competing Interest Statement

The authors have declared no competing interest.

### Author Declarations

The study has been approved by Institutional Review board (IRB) Human Subjects Review Committee of Public Health Institute (IRB#120-002).

