## Supplemental Fig S1 for "Exposome Epidemiology for Suspect Environmental Chemical Exposures during Pregnancy Linked to Subsequent Breast Cancer Diagnosis"

### Supplemental Figure S1

**F1,** <u>4-Hydroxyretinoic acid (315.1957 *m/z*, M-H)</u>

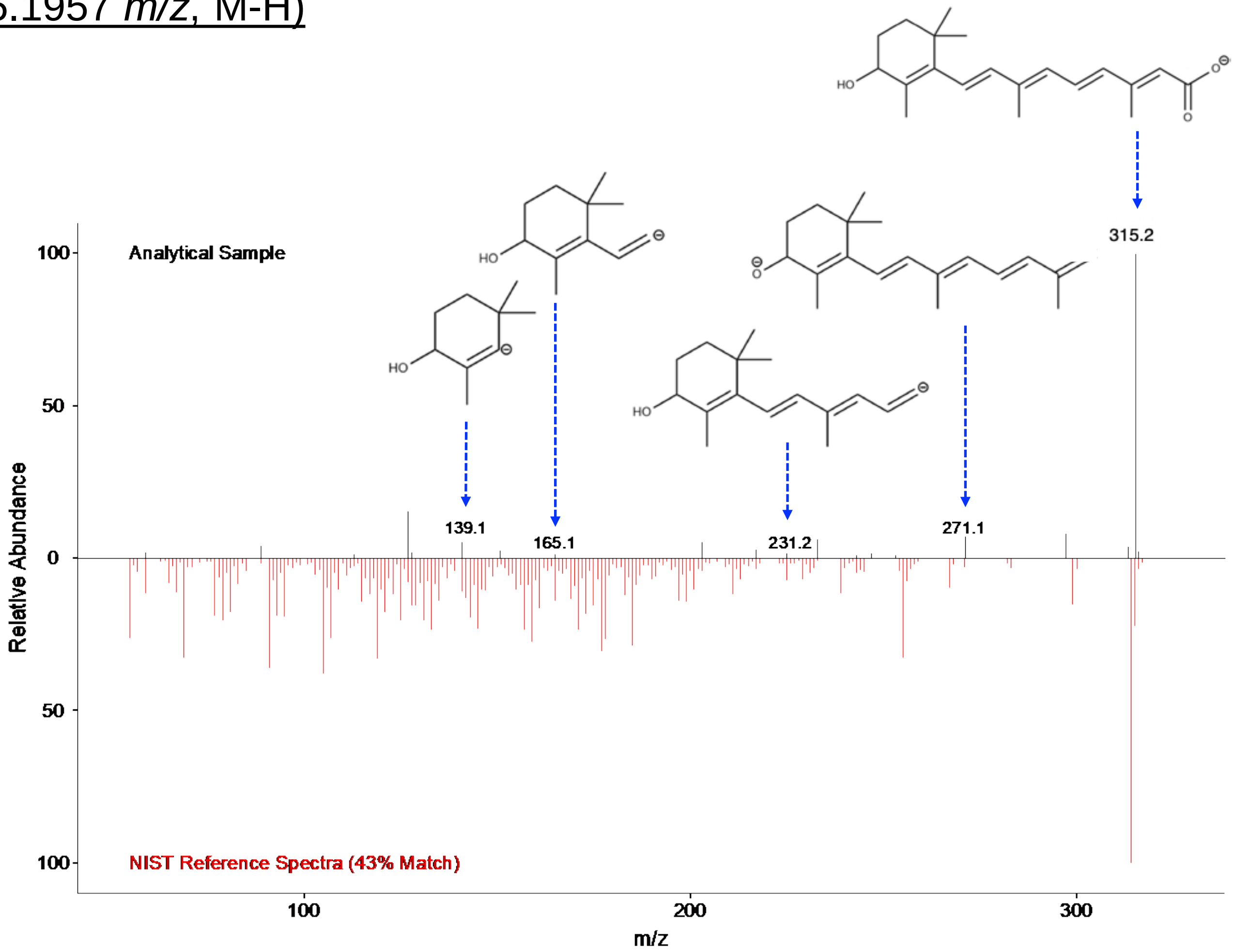


**<u>Reference Spectra</u>**
**Source:** NIST Tandem Mass Spectral Library

### **F3,** 2-Nitromethylene-piperidine (141.067 *m/z*, M-H)

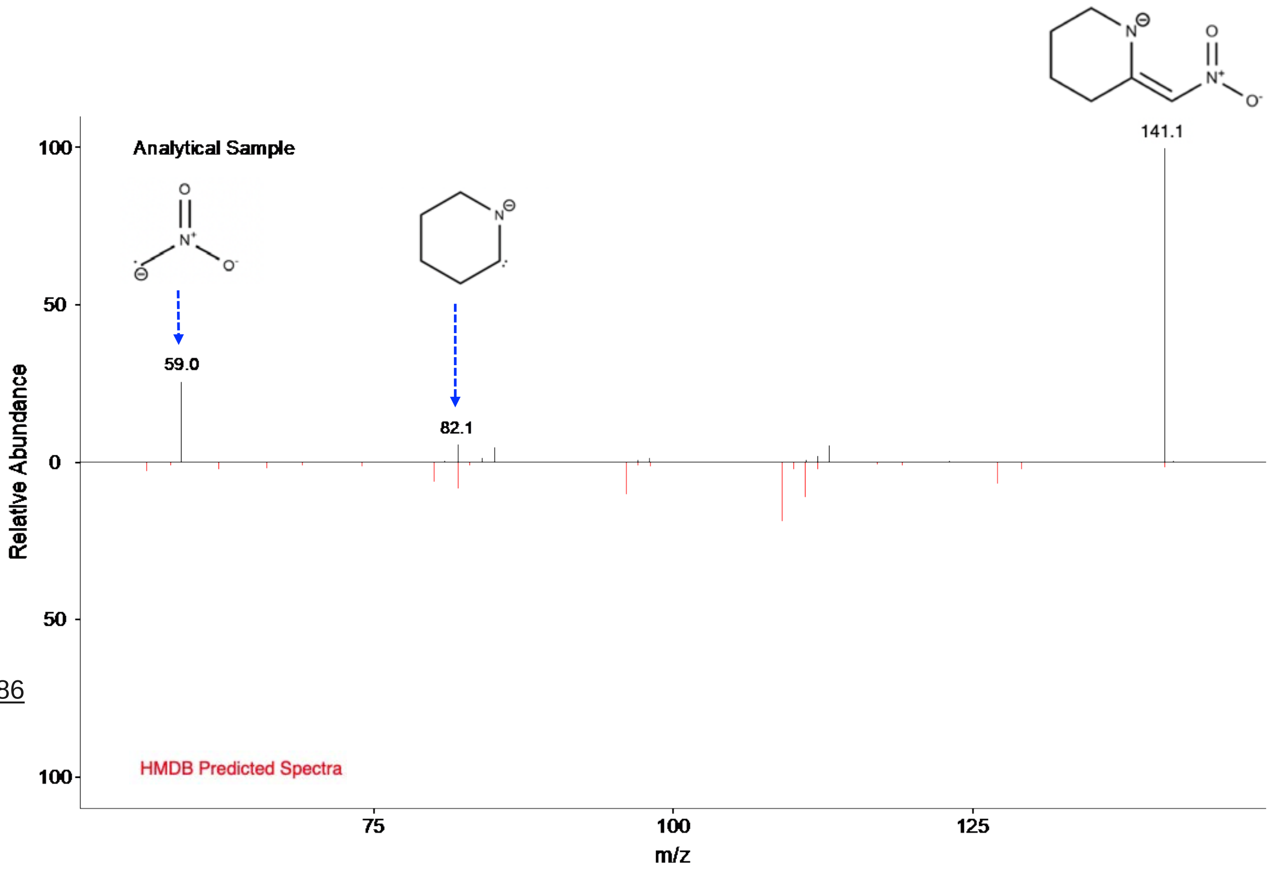


**Reference Spectra**
**Source:** http://www.t3db.ca/spectra/ms_ms/144586
**CE:** -20 V
**Instrument:** LC-MS/MS

**F4,** 2,4-Dinitrophenol (183.0046 *m/z*, M-H)

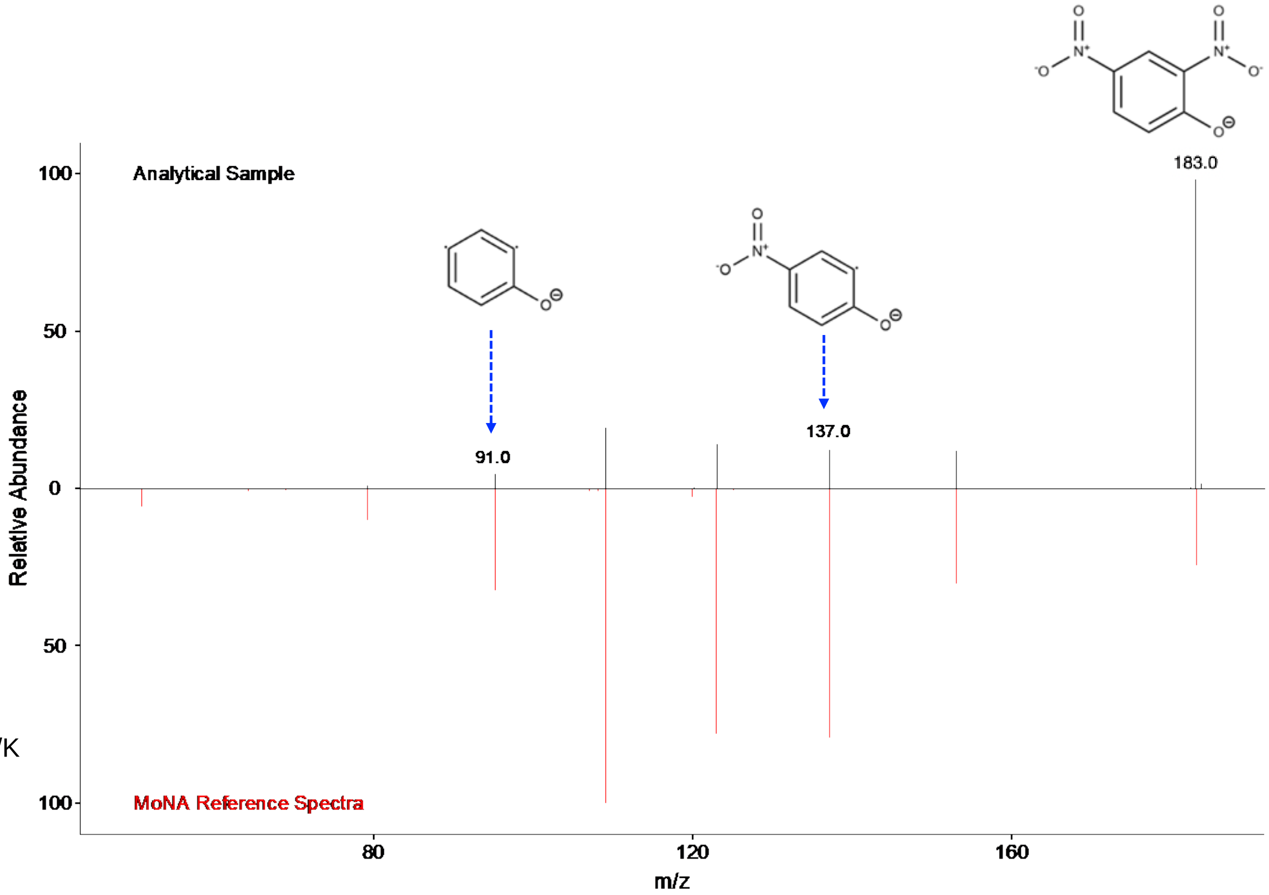


**Reference Spectra**

**Source:** https://mona.fiehnlab.ucdavis.edu/spectra/display/KO000646

**CE:** -30 V

**Instrument:** LC-ESI-QQ

### **F6,** N-Acetylproline (156.0666 *m/z*, M-H)

**Note**
Fragmentation pattern is also consistent with MoNA reference spectra. Source: https://mona.fiehnlab.ucdavis.edu/spectra/display/MoNA023891

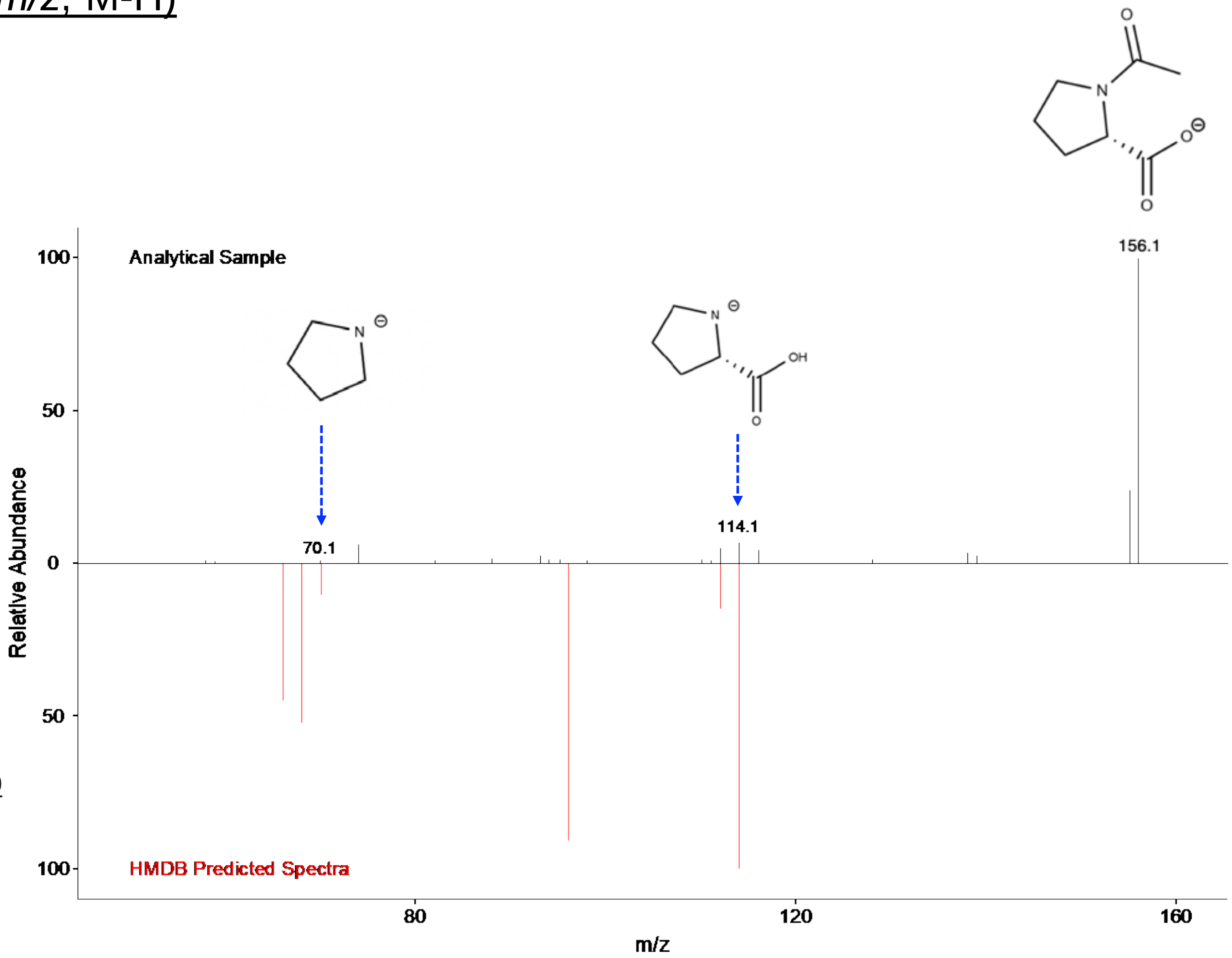


**Predicted Spectra**
**Source:** https://hmdb.ca/spectra/ms_ms/2581210
**CE:** -20 V
**Instrument:** LC-QTOF
