## Supplemental Fig S2 for "Exposome Epidemiology for Suspect Environmental Chemical Exposures during Pregnancy Linked to Subsequent Breast Cancer Diagnosis"

### Supplemental Figure S2

### **F18,** Ergosterol/Ergocalciferol source fragment (271.1195 *m/z*, M-H)

**Note**

Fragmentation of 271.1195 *m/z* showed peaks matching the theoretical structures of ergosterol/ergocalciferol source fragments. The NIST library indicated the fragmentation pattern matched with ergosterol (395.3319 *m/z,* 7% match) and its isomer ergocalciferol (395.3319 *m/z,* 7% match).

**Ergosterol**

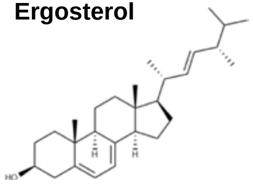

**Ergocalciferol**

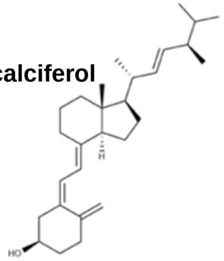

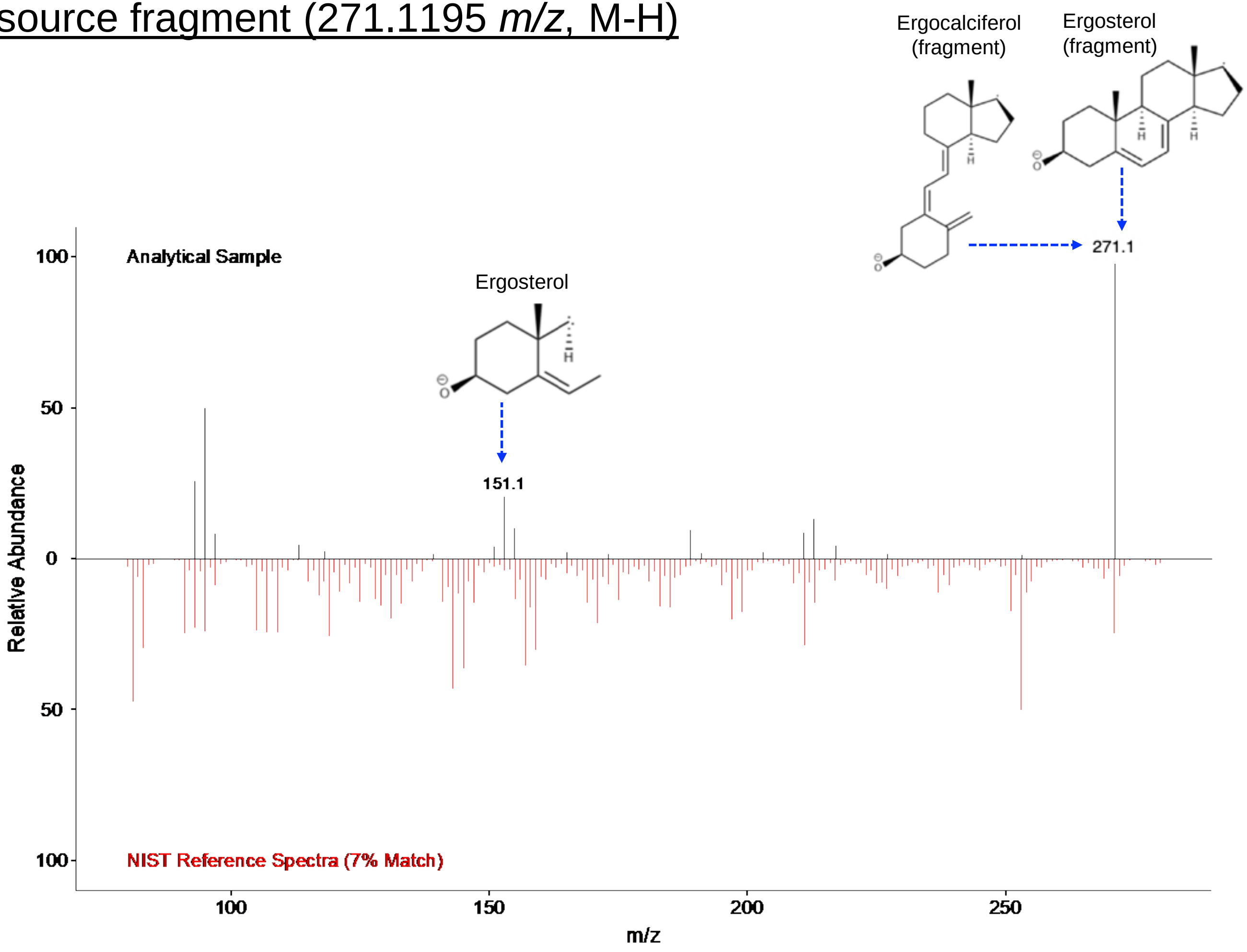


**Predicted Spectra**

**Source:** NIST Tandem Mass Spectral Library

### **F19,** Benzoate derivative (M+Br, 214.9728 *m/z*)

**Note**

All HMDB and T3DB *m/z* matches for 214.9728 *m/z* indicate it is likely a M+Br adduct of a benzoate derivative metabolite. LC data shows 214.9728 *m/z* (+ Br adduct of a benzoate) and a 135.0452 *m/z* (-H adduct of a benzoate) co-elute.

This mass spectrum shows fragmentation of 135.0452 *m/z* and theoretical fragment structures of a common benzoate derivative, 2-methoxybenzaldehyde, compared to its HMDB predicted spectra.

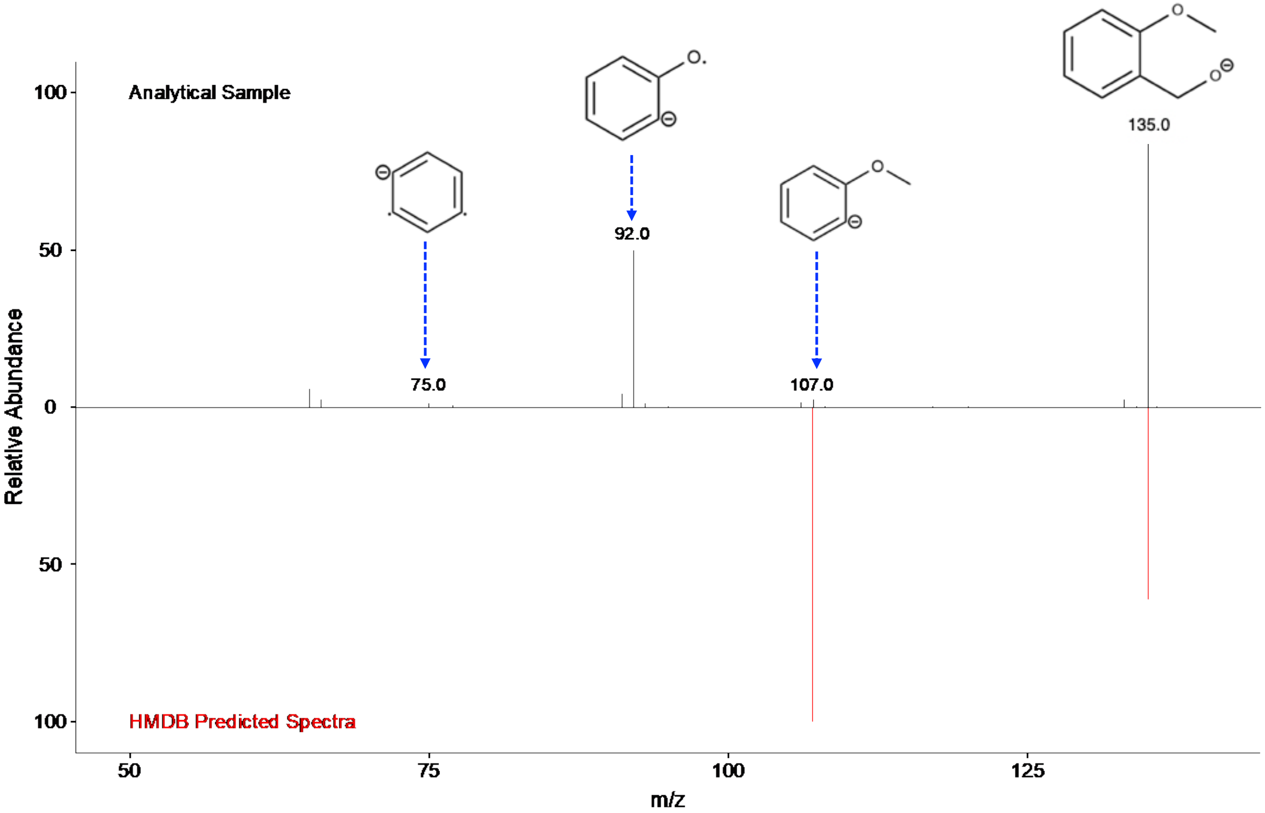


**Predicted Spectra**

**Source:** https://hmdb.ca/spectra/ms_ms/2396004

**CE:** -10 V

**Instrument:** LC-MS/MS

### **F20**, Benzo[a]carbazole, 217.0866 *m/z*

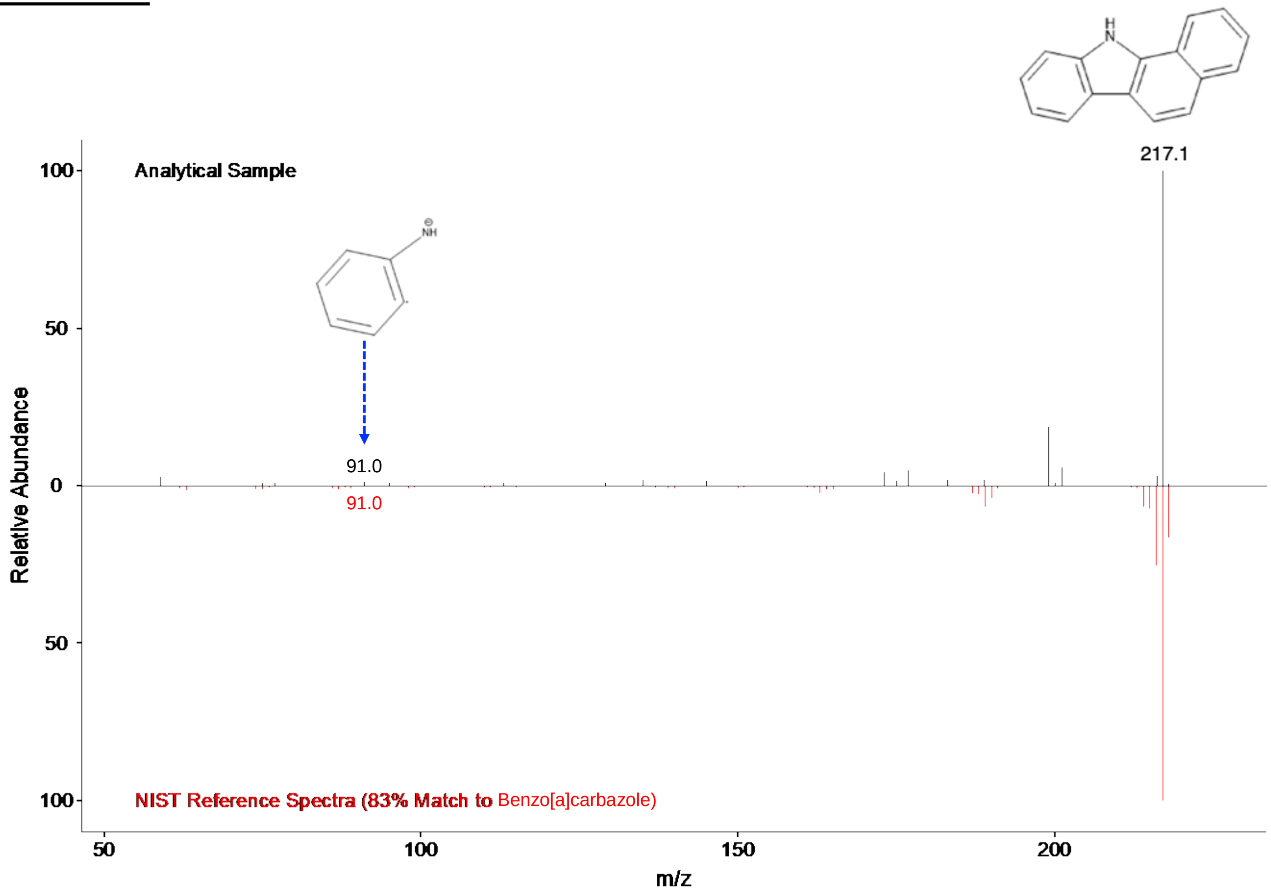


**Reference Spectra**

**Source:** NIST Tandem Mass Spectral Library

### **F21,** Glucose (215.0327 *m/z*, M+Cl$^-$)

**Note**
High signal intensity (E10) and correlates with glucose (M+Na) in HILIC/ESI+ analyses of the same samples.

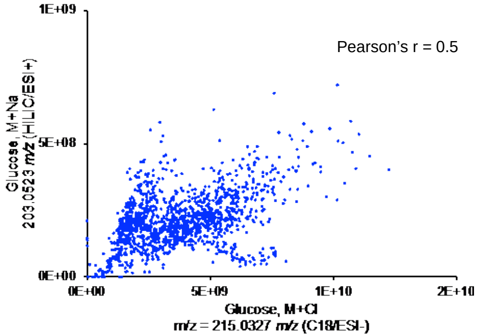


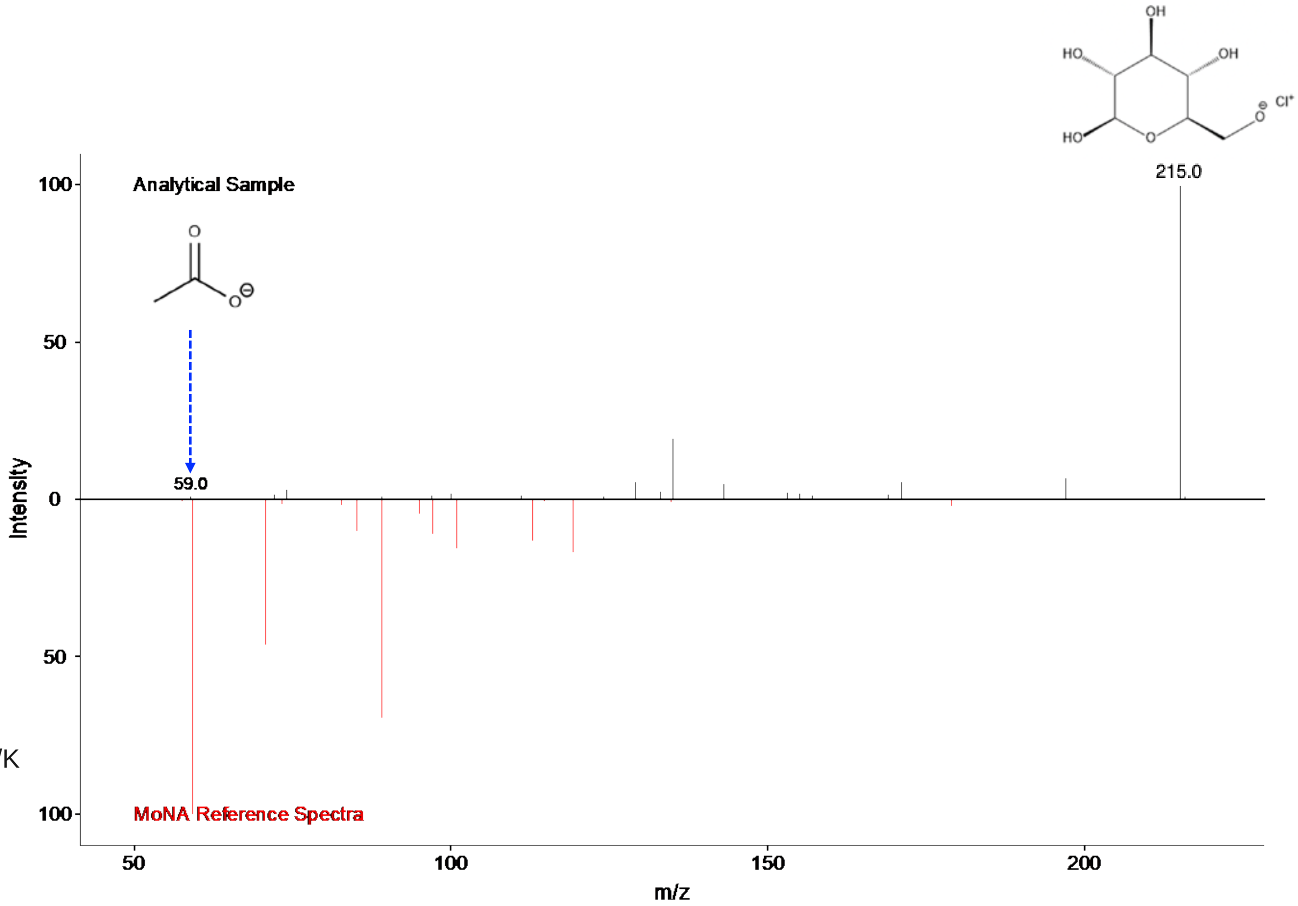


**Reference Spectra**
**Source:**
https://mona.fiehnlab.ucdavis.edu/spectra/display/KO000804
**CE:** -20 V
**Instrument:** LC-ESI-QQ
